# Physical and psychosocial consequences of falls in individuals with cerebral palsy

**DOI:** 10.1101/2023.08.16.23294077

**Authors:** Marissa Thill, Linda E. Krach, Kari Pederson, Nathan G. Wandersee, Sandy Callen Tierney, Elizabeth R. Boyer

## Abstract

**Aim:** To quantify fall frequency, characteristics, injuries, and psychosocial impact across the lifespan and between Gross Motor Classification System (GMFCS) levels in cerebral palsy (CP).

**Method:** Ambulatory individuals with CP (201 adults) or minors’ caregivers (180) completed online surveys on falls and their physical and psychosocial consequences for this cross-sectional study.

**Results:** Almost everyone fell in the past year (86%); GMFCS level II fell most often. Having experienced serious fall-related injuries (e.g., head/face stitches, concussion) increased with age, affecting 80% of ≥50-year-olds. Forward falls caused by tripping during shod ambulation were common. Uneven surfaces and fatigue were notable causes. Concern about falling and associated activity avoidance was highest for GMFCS level III. Psychosocial consequences of falls (e.g., embarrassment, lost confidence) were elevated across GMFCS levels. Nearly everyone (88%) wished they fell less.

**Interpretation:** Fall frequency in ambulatory children and adults with CP is 2-3-fold higher than the general older adult population. Physical and psychosocial consequences of falls were frequent and impacted life choices. Differences observed by GMFCS level and age should be considered in care delivery. Clinically tracking and discussing falls and their repercussions across the lifespan will aid in addressing this under-researched and under-resourced concern of people with CP.

**WHAT THIS PAPER ADDS:**

1. Among 381 individuals with CP, 86% fell in the past year.
2. Falling was most common among 5-12-year-olds and GMFCS level II.
3. Head/face stitches, fractures, sprained knee/ankle, and concussions were common serious injuries.
4. Participants experienced high levels of embarrassment, pain, and loss of confidence.
5. GMFCS level III had the greatest fall concern and avoidance behavior.

## INTRODUCTION

Individuals with cerebral palsy (CP) can experience a variety of sensorimotor impairments that impact balance, making excessive tripping and falling among the most common concerns for ambulatory individuals with CP.^1–3^ Approximately 53-97% of ambulatory individuals fall at least once per year,^4–6^ causing injury, embarrassment, frustration, activity avoidance, and isolation.^6–9^ Although a study of over 1000 children found fall frequency was highest in gross motor function classification system (GMFCS) level II,^4^ research has not explored if this holds across the decades of adulthood. It is unlikely the consequences of falls from these predominantly small, heterogenous samples generalize to all ages and GMFCS levels. Similarly, estimates of fall-related injuries vary greatly (31-82%)^6,8^ and should be examined by age and GMFCS level.

Furthermore, there is limited evidence of the psychosocial consequences of falls.^6–11^ Concern about or fear of falling is potentially more disabling than the fall itself. Embarrassment, frustration, injury, pain, and feelings of isolation contribute to concern about falling and subsequent activity avoidance.^6,7,9^ Because individuals with CP across all ages are less physically active^12,13^ and are at higher risk for sedentary-related conditions,^14,15^ it is imperative their balance impairment and fall anxiety is managed so it does not perpetuate decreased physical activity, participation, or wellbeing.^16^ The purpose of this survey-only study was to address these gaps by quantifying fall frequency, characteristics, and consequences (physical and psychosocial) in individuals with CP, stratified by age and GMFCS level.

## METHODS

The University of Minnesota Institutional Review Board approved this study. The inclusion criteria were individuals with CP (or minors’ caregiver), aged 5-89, GMFCS levels I-III, reads English, and their own legal decision-maker (adult participants only).

Recruitment occurred via Gillette Children’s patient emails (visits from 2011-2021) and social media listings, as well as Cerebral Palsy Research Network’s community registry (25-89-year-olds only).

Research Electronic Data Capture (REDCap) was used for screening, consent, and surveys (Table 1). Caregivers were asked to consult their child, as appropriate. Surveys included (1) fall history (serious and minor injuries), (2) fall characteristics (frequency in the past 1 and 12 months; direction; cause; use of assistance; foot wear), (3) Short Falls Efficacy Scale-International (FES-I), (4) Short FES-I Avoidance Behavior (FES-IAB),^17^ and Consequences of Falling-Damage to Identity.^18^ For Damage to Identity, participants were prompted to think about falls in public. Four optional open-ended falls questions were included.

**Table 1.**
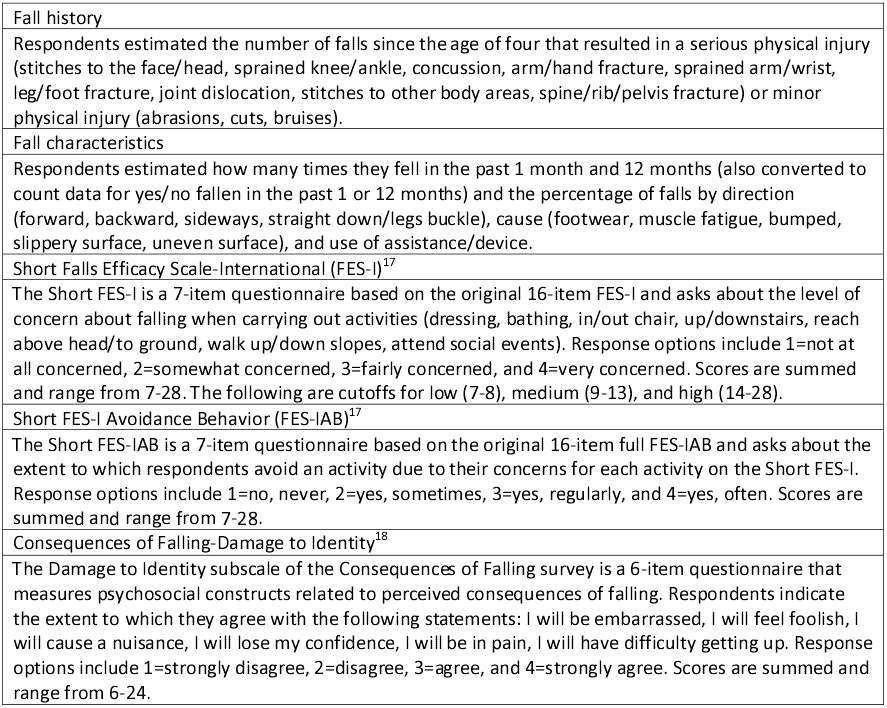
Brief description of the study surveys.

### Data Analysis

Missing data were excluded. All continuous data were non-parametric, so main effects for age (groups: 5-12, 13-17, 18-34, 35-49, ≥50) and GMFCS level were assessed using two Kruskal-Wallis ANOVAs. For count data, chi-square tests were used to assess age and GMFCS level effects. If main effects were present, Tukey-adjusted pairwise comparisons were performed.^19^ Analyses were performed in Matlab (R2021b, Natick, MA, USA). Significance was set at p<0.05.

Framework Theory with deductive and inductive approaches guided content analysis of the open-ended questions to create initial and final codes and a codebook.^20^ Responses were analyzed by two authors (MT, ERB). People with lived experience informed the codebook and joined disagreement discussions to help reach consensus.

## RESULTS

The final sample included 381 participants (201 adults; Table 2). Expanded results are provided elsewhere (Supporting Information). Quantitative results are the focus of this paper.

**Table 2.** Participant demographics.

| GMFCS level | Age group (y) | Age (y), mean (SD) [min-max] | n | Male, n (%) | Topography, n (%) |  |  |  |  | Cognitive impairment,* n (%) |
| --- | --- | --- | --- | --- | --- | --- | --- | --- | --- | --- |
|  |  |  |  |  | Mono/Hemi | Di | Tri | Quad | Other/Unk |  |
| all |  | **24 (17) [5-76]** | **381** | **177 (46%)** | **142 (37%)** | **111 (29%)** | **39 (10%)** | **77 (20%)** | **12 (3%)** | **119 (31%)** |
| I | all | **22 (15) [5-71]** | **126** | **66 (53%)** | **78 (62%)** | **23 (18%)** | **10 (8%)** | **13 (10%)** | **2 (2%)** | **28 (22%)** |
|  | 5-12 |  | 39 | 23 (61%) | 30 (77%) | 4 (10%) | 4 (10%) | 1 (3%) | 0 | 9 (23%) |
|  | 13-17 |  | 25 | 16 (64%) | 17 (68%) | 1 (4%) | 3 (12%) | 4 (16%) | 0 | 11 (44%) |
|  | 18-34 |  | 42 | 18 (43%) | 24 (57%) | 10 (24%) | 2 (5%) | 4 (10%) | 2 (5%) | 8 (19%) |
|  | 35-49 |  | 9 | 3 (33%) | 3 (33%) | 4 (44%) | 0 | 2 (22%) | 0 | 0 |
|  | ≥50 |  | 11 | 6 (55%) | 4 (36%) | 4 (36%) | 1 (9%) | 2 (18%) | 0 | 0 |
| II | all | **25 (18) [5-76]** | **168** | **72 (44%)** | **53 (32%)** | **59 (35%)** | **15 (9%)** | **36 (21%)** | **5 (3%)** | **57 (34%)** |
|  | 5-12 |  | 50 | 31 (63%) | 23 (46%) | 13 (26%) | 4 (8%) | 8 (16%) | 2 (4%) | 30 (60%) |
|  | 13-17 |  | 27 | 13 (50%) | 7 (26%) | 8 (30%) | 6 (22%) | 3 (11%) | 3 (11%) | 12 (44%) |
|  | 18-34 |  | 50 | 17 (36%) | 12 (24%) | 19 (38%) | 4 (8%) | 15 (30%) | 0 | 15 (30%) |
|  | 35-49 |  | 17 | 3 (19%) | 5 (29%) | 7 (41%) | 1 (6%) | 4 (24%) | 0 | 0 |
|  | ≥50 |  | 24 | 8 (33%) | 6 (25%) | 12 (50%) | 0 | 6 (25%) | 0 | 0 |
| III | all | **25 (18) [5-70]** | **87** | **39 (46%)** | **11 (13%)** | **29 (33%)** | **14 (16%)** | **28 (32%)** | **5 (6%)** | **34 (40%)** |
|  | 5-12 |  | 28 | 14 (52%) | 4 (14%) | 10 (36%) | 7 (25%) | 6 (21%) | 1 (4%) | 11 (41%) |
|  | 13-17 |  | 11 | 4 (40%) | 0 | 4 (36%) | 3 (27%) | 4 (36%) | 0 | 7 (64%) |
|  | 18-34 |  | 29 | 15 (54%) | 4 (13%) | 8 (28%) | 1 (3%) | 13 (45%) | 3 (10%) | 14 (48%) |
|  | 35-49 |  | 8 | 3 (38%) | 2 (25%) | 2 (25%) | 1 (13%) | 3 (38%) | 0 | 2 (25%) |
|  | ≥50 |  | 11 | 3 (27%) | 1 (9%) | 5 (45%) | 2 (18%) | 2 (18%) | 1 (9%) | 0 |

\* self-reported as at least mild cognitive impairment
Di: diplegia, Hemi: hemiplegia, Mono: monoplegia, SD: standard deviation, Tri: triplegia, Unk: unknown/not reported, y: years

### Fall Frequency and Characteristics

Eighty-six percent (standard error (SE): 2%) reported at least one fall last year—89% of children, 85% of adults. The youngest group, ages 5-12, fell most frequently in the past month (χ^2^=39.0, p<0.001) and year (χ^2^=36.6, p<0.001, Figure 1). Monthly and yearly falls differed by GMFCS level (χ^2^=11.8, p=0.003 and χ^2^=7.7, p=0.021); GMFCS level II fell more (median: 8 falls/year) than level I (4) but not level III (6).

**Figure 1.**
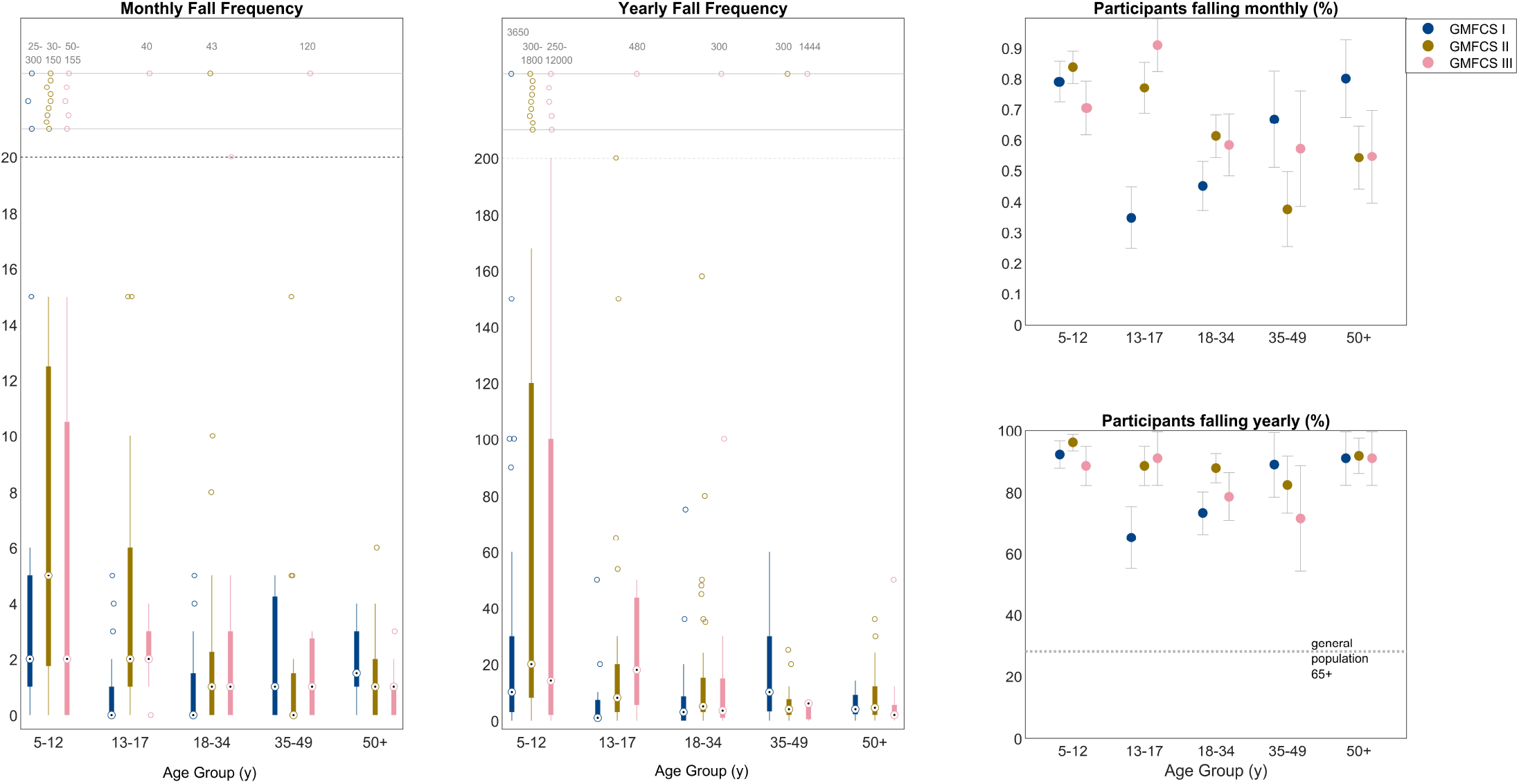
Frequency of falls in the past 1 month (left), 12 months (middle), and proportion of participants falling in the past 1 month (top right) or 12 months (bottom right) by GMFCS level and age group. (Left & Middle) The median is represented as the bullseye and 25-75%tile as the edge of the boxes. Outliers are plotted as ‘o’. Extreme outliers were plotted above the black dashed line with numerical range of extreme outliers given. (Right) Error bars represent the standard error of the estimate. The horizontal dotted line represents 28% of US adults aged ≥65 years who report falling in the past 12 months. 21GMFCS: Gross Motor Function Classification System, y: years.

Fall characteristics were similar across age (except the oldest wore shoes more; the youngest were bumped more; Supporting Information), though characteristics often differed by GMFCS level (Table 3). Overall, participants estimated nearly all falls were during shod ambulation in the forward direction. Uneven surfaces and fatigue were common causes.

**Table 3.** The proportion (median [IQR]%) of falls associated with the various fall characteristics and main effects by age* and GMFCS level. If a main effect was present, pairwise comparisons were investigated, and significant differences reported.

|  | ALL | GMFCS I | GMFCS II | GMFCS III | Age effect | GMFCS effect |
| --- | --- | --- | --- | --- | --- | --- |
| **Falls wearing shoes** | 50 [20-89] | 50 [10-90] | 50 [20-85] | 50 [15-80] | $\chi^2=19.6$, $p<0.001$, Differences: 5-12 vs. $\geq 50$, 13-34 vs. $\geq 50$ | $\chi^2=0.2$, $p=0.914$ |
| **Falls wearing socks only** | 10 [0-25] | 10 [0-20] | 10 [2-25] | 5 [0-21] | $\chi^2=13.6$, $p=0.009$, Differences: 5-12 vs. 35-49, 5-12 vs. $\geq 50$ | $\chi^2=5.4$, $p=0.068$ |
| **Falls while barefoot** | 10 [1-30] | 10 [0-20] | 15 [5-35] | 15 [0-40] | $\chi^2=16.5$, $p=0.003$, Differences: 5-12 vs. 35-49, 5-12 vs. $\geq 50$ | $\chi^2=12.3$, $p=0.002$, Differences: I vs. II |
| **Falls during ambulation** | 85 [50-100] | 90 [55-100] | 90 [50-100] | 75 [25-100] | $\chi^2=7.0$, $p=0.132$ | $\chi^2=6.7$, $p=0.035$ Differences: I vs. III |
| **Falls using assistance (device or person)** | 2 [0-16] | 0 [0-5] | 3 [0-10] | 10 [1-75] | $\chi^2=6.0$, $p=0.196$ | $\chi^2=45.6$, $p<0.001$, Differences: all groups |
| **Forward falls** | 65 [33-90] | 80 [45-99] | 70 [40-90] | 40 [18-75] | $\chi^2=4.3$, $p=0.363$ | $\chi^2=18.2$, $p<0.001$. Differences: I vs. III, II vs. III |
| **Backwards falls** | 10 [1-25] | 7 [0-20] | 10 [2-25] | 25 [5-46] | $\chi^2=2.9$, $p=0.583$ | $\chi^2=22.6$, $p<0.001$ Differences: I vs. III, II vs. III |
| **Sideways falls** | 10 [0-25] | 5 [0-20] | 10 [2-25] | 10 [5-25] | $\chi^2=5.8$, $p=0.214$ | $\chi^2=6.0$, $p=0.050$, Differences: none |
| **Downward falls** | 2 [0-20] | 0 [0-10] | 2 [0-15] | 10 [0-25] | $\chi^2=3.5$, $p=0.471$ | $\chi^2=12.4$, $p=0.002$, Differences: I vs. III |
| **Falls on uneven surfaces/stairs** | 39 [15-60] | 50 [15-70] | 40 [20-63] | 25 [10-50] | $\chi^2=3.7$, $p=0.443$ | $\chi^2=9.3$, $p=0.010$, Differences: I vs. III, II vs. III |
| **Falls caused by fatigue** | 25 [5-50] | 20 [5-50] | 25 [10-50] | 25 [5-60] | $\chi^2=4.2$, $p=0.375$ | $\chi^2=2.2$, $p=0.342$ |
| **Falls caused by being bumped** | 15 [5-30] | 10 [3-25] | 20 [10-30] | 20 [10-40] | $\chi^2=18.5$, $p=0.001$, Differences: 5-12 vs. 18-34 | $\chi^2=6.2$, $p=0.0453$, Differences: none |
| **Falls caused by slippery surface** | 15 [5-30] | 15 [5-30] | 20 [5-30] | 10 [5-25] | $\chi^2=2.8$, $p=0.587$ | $\chi^2=2.1$, $p=0.343$ |

\*Results by age groups are presented in Supporting Information.
IQR: interquartile range (25-75<sup>th</sup> percentile), vs: versus

### Physical Consequences

Across participants, 55(3)% reported at least one fall causing serious injury (Figure 2), though this varied by age group (40(5)%, 44(6)%, 59(5)%, 71(8)%, 80(6)%; χ^2^=29.5, p<0.001). The oldest group experienced more serious injuries than the three youngest groups (>50% of the oldest GMFCS level II-IIIs experienced ≥3; Supporting Information). The main effect for GMFCS was insignificant (χ^2^=4.6,p=0.099), though the 5-12-year-old GMFCS level II group reported nearly twice as many falls resulting in serious injuries (56(7)%) as GMFCS level I or III 5-12-year-olds. Stitches on the head were the most common serious injury (119 reported; 22% of participants), followed by sprained knee/ankle (89; 16%) and concussions (78; 14%). Overall, 20(2)% had at least one fracture, affecting about 40% of ≥50-year-olds (Supporting Information). Other self-reported serious injuries included herniated discs and broken teeth.

**Figure 2.**
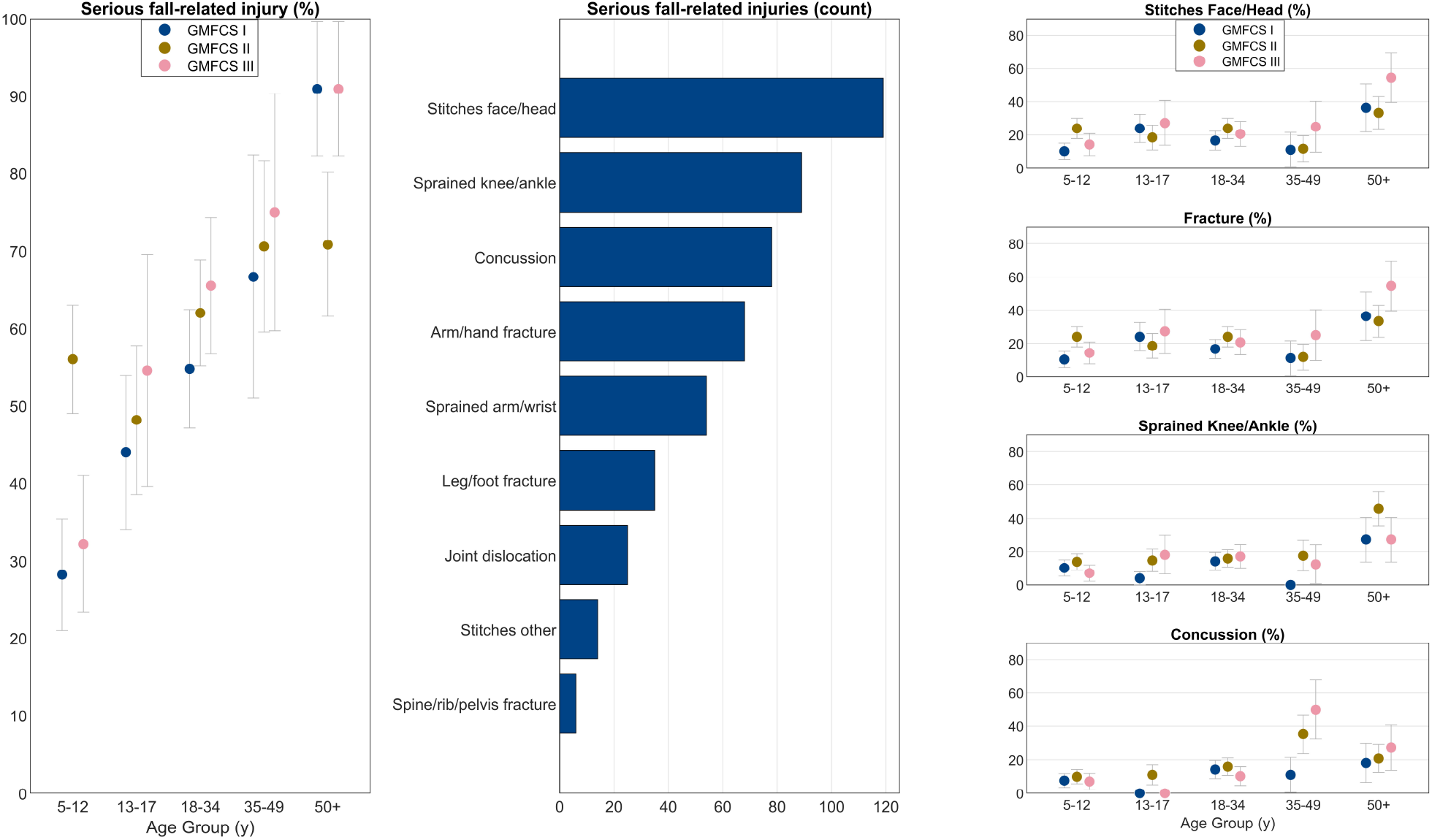
Serious fall-related injuries. (Left) Proportion of respondents reporting 1 or more fall that resulted in serious injury(ies) by GMFCS level and age group. Error bars represent the standard error of the estimate. (Middle) Count of serious fall-related injury types among the entire cohort. (Right) Proportion of respondents who had the four most common serious fall-related injuries by GMFCS level and age group (any fracture was combined here to include: shoulder, arm, wrist, hand, spine, ribs, pelvis, leg, or foot). Error bars represent the standard error of the estimate. GMFCS: Gross Motor Function Classification System, y: years.

Minor fall-related injuries (abrasions, cuts, bruises) were more common to the body (median: 20, range: 0-1500) than face (1, 0-300). Age effects were noted for the number of minor facial (χ^2^=12.2, p=0.016) and bodily injuries (χ^2^=9.9, p=0.042). Ages 5-12 had significantly more facial injuries than ages 13-17. Ages 35-49 had significantly more bodily injuries than 5-12 and 13-17-year-olds. There was no difference by GMFCS level for minor facial (χ^2^=2.5, p=0.293) or bodily injuries (χ^2^=5.7, p=0.057; median: 20).

Median ‘pain’ scores (from Damage to Identity) for each age-GMFCS group were 3-3.5 (3: agree, 4: strongly agree). Median ‘difficulty getting up’ ranged from 2 (disagree) for the three youngest GMFCS level I groups, 3 for most others, and 4 for the oldest GMFCS level III group.

### Psychosocial Consequences

Short FES-I differed by age (χ^2^=13.5, p=0.009) and GMFCS (χ^2^=103.2, p<0.001, Figure 3, Supporting Information). Concern about falling was significantly greater in ≥50-year-olds versus 18-34-year-olds. As motor impairment (GMFCS level) increased, so did concern about falling (all pairwise comparisons were significant). Short FES-IAB differed by age (χ^2^=19.2, p<0.001) and GMFCS (χ^2^=70.0, p<0.001). The ≥50-year-olds showed more avoidance than the three youngest groups. All GMFCS levels differed from each other; as motor impairment increased, so did avoidance. Fall concern and avoidance tended to be highest on slopes and stairs.

**Figure 3.**
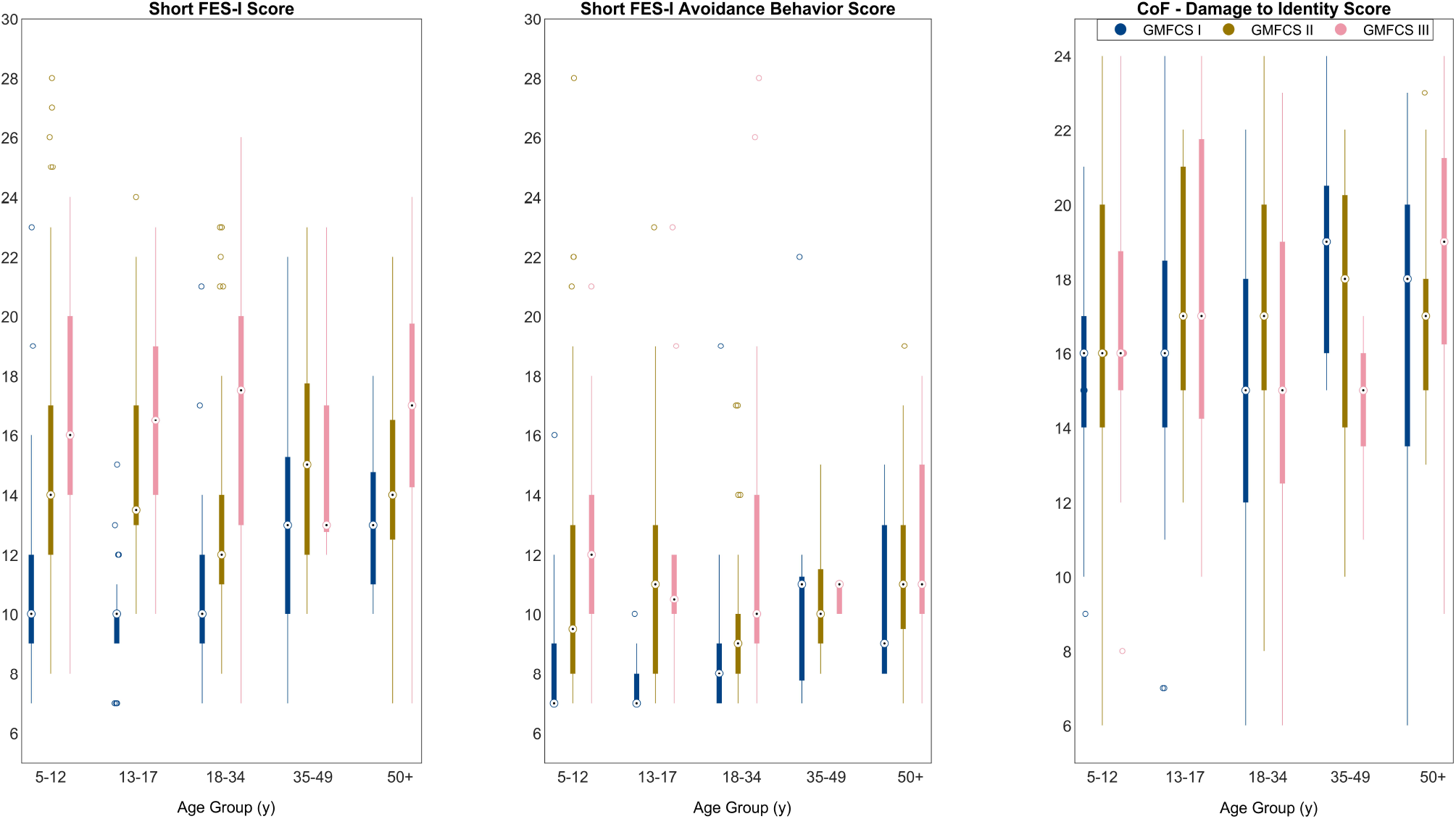
Scores for concern about falling (Left), avoidance behavior due to concern about falling (Middle), and psychosocial impact of falls (Right) by GMFCS level and age group. The median is represented as the bullseye and 25-75%tile as the edge of the boxes. Outliers are plotted as ‘o’. Since possible score ranges differ between the Short FES-Is and Damage to Identity, scores should not be compared across surveys. CoF: Consequences of Falling, FES-I: Falls Efficacy Scale-International, GMFCS: Gross Motor Function Classification System, y: years.

Damage to Identity did not differ by age (χ^2^=7.1, p=0.130) or GMFCS (χ^2^=5.0, p=0.083). Embarrassment, pain, and losing one’s confidence were scored the highest of the six items.

Nearly all participants wished they fell less (estimate(SE): 88(2)%). There was no difference by age (χ^2^=5.1, p=0.279) but there was by GMFCS level (χ^2^=19.4, p<0.001), with fewer GMFCS level I participants (77(4)%) reporting this sentiment than level II (94(2)%) and III (90(3)%).

### Qualitative Results

Two central themes (internal, external) and eight subthemes emerged (Table 4). Participants elaborated on fall causes, mechanics, repercussions, frustrations, and desires for themselves and society.

**Table 4.**
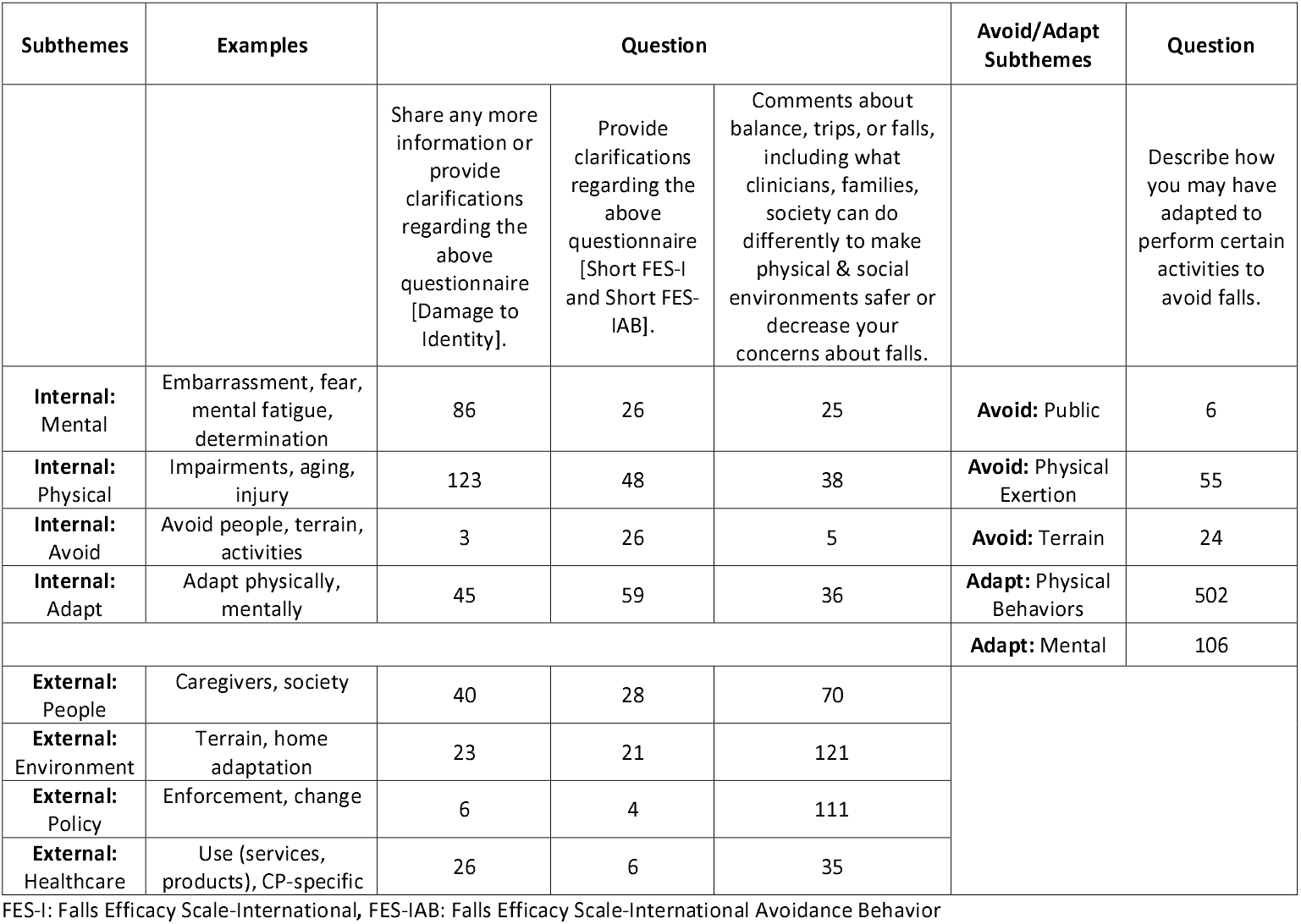
Number of comments pertaining to subthemes that emerged from the qualitative analysis of the four optional open-ended questions.

Causes included physical, mental, environmental, and situational factors (e.g., muscle fatigue, aging, uneven surface). Participants often cited foot clearance issues (vs. slips), though falls also occurred during transfers for individuals with greater motor impairment.

> *“The older I get, the less confident I am in my balance and the slower my reaction times become*.*”*
>
> *“If I get nervous then my muscles tense up, and it takes a bit for them to calm down*.*”*
>
> *“…mental fatigue/being distracted is the cause of the overwhelming majority of my falls. If I’m not THINKING enough to pick up my foot, BAM*.*”*

Physical repercussions were present and changed with time, including difficulties getting up.

> *“…the severity of my injuries grew as I reached middle age and my muscles naturally shortened*.*”*

Psychosocial repercussions were vast, with a glimpse of how perspectives, like resilience or despondence, can impact participation and quality of life. Planning how to be safe was common.

> *“I am more concerned about the embarrassment of falling rather than the potential physical damage*.*”*
>
> *“…there will always be hazards…learn approaches to keep yourself safe. “*
>
> *“Sometimes I just sit there and watch everyone else have fun, because I hate my disease*.*” “I worry they will stop seeing me as an equal and more as a liability*.*”*
>
> *“It lowers confidence, increases fear and [my child does] not want to go out and access life as a result. Especially after a hard fall*.*”*

These fall experiences play a role in how participants adapt or avoid activities, including sitting down to dress/shower, crawling/scooting, wearing better shoes, and using assistive devices, braces/orthoses, or people for support.

> “*Am very careful as I walk and limit distances when I’m fatigued*…*”*
>
> *“Walking down [slopes] can be problematic because of my balance, so I generally either ask for help or avoid it*.*”*

Participants’ frustrations and desires addressed public behavior (do not rush, crowd, or overreact), environment (better lighting; declutter; fix uneven surfaces; fewer slopes; more handrails/grab bars, benches, curb cuts; timely removal of snow/deicing), policy (enforce the Americans with Disabilities Act), and healthcare (outdoor therapy, lifelong therapy, insurance coverage).

> *“Ask me if I need help getting up…and how I prefer to be assisted*.*” “Ban scatter rugs*.*”*
>
> *“Teach kids how to fall in a way that minimizes injuries*.*”*
>
> *“Have Medicare allow wheelchairs for the longer [distances]*.*”*
>
> *“Please stop providing patients with lifelong disabilities the same fall risk advice you provide elderly patients*.*”*

## DISCUSSION

This novel study reported fall frequency, characteristics, and consequences across the lifespan for ambulatory individuals with CP. Fall rates and physical and psychosocial consequences were universally high compared to the general population, with individuals in GMFCS levels II and III being most affected by falls. The most salient difference by age was more frequent falls in childhood than adulthood.

Over 80% of participants fell in the past year, representing the higher end of the range reported in adult CP studies.^4–6^ This is three times higher than the general older adult population (28%)^21^ and exceeds that of multiple sclerosis, Parkinson’s Disease, amputees, and stroke (∼60%).^22–25^ Similar to previous findings,^4,26^ individuals in GMFCS level II fell most often, as they have greater impairments than individuals in level I but do not frequently use assistive devices like level III. The number of falls varied across groups, with the youngest group falling most often (medians: 10-20), some >1000 times/year. This may be expected, as even novice walkers (age 12-19 months) without disabilities fall about 17 times/hour.^27^ Some adults in GMFCS level II and III reported >100 falls/year, which concurs with others,^26^ though the groups’ medians were <10 in this study. It is advisable that clinicians share this information with families to help reassure them of the commonality of falls and that falls generally decrease through middle adulthood. Larger samples sizes are needed to understand trends beyond that.

How participants fell and the causes tended to differ more by GMFCS level than age. Notably, individuals in GMFCS level III were less likely to fall forward, during ambulation, and on uneven surfaces but more likely to fall down/legs buckle compared to other participants. This is likely due to greater impairment of their strength, endurance, motor control, and balance. Despite this difference by GMFCS level, ≥75% of all falls occurred during ambulation, often on uneven surfaces, agreeing with other reports.^6^ Slippery socks were rarely the culprit of falls. While fatigue is a common concern in CP,^1^ participants estimated only 25% of their falls were due to fatigue. Researchers and therapists would be prudent to incorporate these findings into study designs and care plans.

While many participants reported the ability to stand up unscathed after most falls, falls can have several physical consequences, including injury, pain, difficulty getting up, and sedentary behavior due to activity restriction. Over half of participants reported a fall causing serious injury(ies). The most common but least serious of these injuries, facial stitches, may be embarrassing due to their conspicuousness. Serious injuries, like fractures and sprains, can result in multi-week activity restriction and disruption of life responsibilities. Prolonged activity restriction, per medical advice and/or cautiousness/anxiety, can decrease fitness and bone health, further placing participants at risk of fractures and sedentary-related conditions. It is understandable that many adults with CP attribute their mobility decline to impaired balance and falls.^10,16^ Still, other long-term health ramifications can result. One 5-12-year-old had eight concussions, making them 1 of 13 participants (3%) who had more than one concussion. These high concussion rates are worrisome since long-term effects include depression, anxiety, and cognitive issues, which are already high in individuals with CP.^28,29^ These factors may create a vicious cycle of self-perpetuating health challenges, magnified by inevitable aging effects.

Both quantitative and qualitative data revealed elevated concerns about falling and associated negative emotions, such as embarrassment, foolishness, despondence, loss of confidence, and activity avoidance, which have been observed elsewhere.^6–9^ Short FES-I scores for GMFCS levels II and III herein were higher than the general older adult population (medians: 12-17 versus 12) and within the moderate-to-high range reported by adults with CP.^8,10,11^ FES-I has not been measured in children with CP. This study’s results reveal a chronic, elevated concern about falling since childhood that transforms into anxiety for some and takes a toll on oneself/caregivers and behavior choices. We observed a difference in Short FES-I between the youngest and oldest adults, which may be driven by decreased balance, strength, and reaction times with age, recounted by several participants. A 35-49-year-old participant shared that they worry about falling more now than before. They intentionally avoid high-risk situations since an injury could impede their ability to provide for their family as the primary household earner. These avoidance behaviors were evident on a group level. Short FES-IAB scores were largest for the older group compared to the three youngest groups. While median scores were relatively low, they were higher than the general older adult population.^17^

Damage to Identity scores showed no differences by age or GMFCS level, even though age and GMFCS were associated with differences in fall frequency, serious fall-related injuries, and concern about falling.

Scores were universally high, though there was a trend toward older groups reporting greater psychosocial impact, consistent with other studies.^6,9^ While scores herein exceeded typical ≥75-year-olds’,^18^ the difference may be due to survey modification. During study planning, people with lived experience said embarrassment/foolishness would differ based on who witnessed the fall, so we modified the survey instructions to consider public falls. Nonetheless, embarrassment came up repeatedly in the qualitative component. The inevitability of falls, fearfulness, and embarrassment have been shown here and by others to impact the ability of some individuals to fully participate in life.^6,7,9^

This study generated useful information and calls for action beyond the person with CP or caregiver. Age- and GMFCS-specific results can help caregivers and clinicians identify children at disproportionately higher risk of falls and associated outcomes and anticipate their changes with age. Keeping a fall diary may be beneficial to share with clinicians. International, multi-stakeholder efforts recently produced a core outcome set for lower limb orthopedic surgery, which includes balance and falls;^30^ identifying appropriate assessments is the logical next step. Inpatient fall risk assessments (e.g., Humpty Dumpty, Morse) are inadequate for this purpose since they focus on inpatient risk. Since reporting falls may feel like it jeopardizes one’s independence, trust is essential. Still, falls are inevitable. Teaching individuals to fall safely and how to get up is critical, which our results suggest is especially important for older individuals in GMFCS level III. Having balance training covered by healthcare insurance across the lifespan, including prophylactically, was advocated, as was CP-specific fall prevention education. Equally important is the identification of at-risk individuals for mental health concerns due to fall experiences, as these effects may compound with time if not addressed. Making timely referrals to address anxieties or avoidance behaviors may improve mental and physical health, as highlighted by participants.

Active research in CP on real-world fall causes, automated fall-detection, and effectiveness of various interventions or devices is warranted. The nearly ubiquitous desire to fall less is reason enough. Study conditions should emulate fall-risk conditions (uneven surface, tripping, walking, fatigue). Investigating the impact of falls on economics (healthcare system, lost wages for affected person) and quality of life is warranted to further justify policy change and enforcement.

The results include a call to action for the public at the individual and policy level. Participants identified myriad helpful responses: showing empathy, giving space, slowing down, not overreacting to falls, asking if/how to help, removing trip hazards, ensuring universal design and inclusive activities (especially at schools), and voting for and enforcing disability policies. These supportive actions foster a safe environment, mitigate the psychosocial distress of falls, and encourage participation rather than avoidance.^7,9^

Despite many novel findings, study limitations should be acknowledged. Small sample sizes (n<20 for 6/15 age-GMFCS groups) decreased statistical power and estimate precision (e.g., percentage of serious injuries did not always logically plateau/increase with age). Second, the results might not generalize if a disproportionate number of people who fell frequently or experienced injuries participated. Third, fall and injury recall was challenging in this cross-sectional study as noted by several participants. For example, the maximum estimate for annual falls (12,000; about 33/day; participant age <10, GMFCS III) exceeded the next highest of 3650 (age <10, GMFCS I), so we are unsure of the veracity of everyone’s estimates. However, the outliers were the youngest participants and may reflect natural, but delayed, development.^27^ Prospective studies are needed, ideally with passive fall detection methods. Fourth, less than half of caregivers consulted with their child (24% for children <8 years; 47% for 8-17 years); therefore, it remains equivocal whether FES-I, FES-IAB, and Damage to Identity responses are projections of the caregiver or representative of minors. Lastly, a design flaw resulted in participants who had a minor facial injury being the only participants asked about minor bodily injuries (n=159), so those results may not generalize.

## Conclusion

Balance and falls have widespread effects on the life of people with CP, highlighting the need to measure fall outcomes in clinics and research. Fall frequency and physical and psychosocial sequelae are disproportionately borne by certain age and GMFCS levels. This knowledge can help tailor falls prevention and care, while also reassuring people with CP that they are not alone in these experiences. Alternatively, some psychosocial consequences did not differ by age or GMFCS level, though scores were universally high compared to the general population. Resources are justified to address this population’s fall-related concerns.

## Data Availability

All data produced in the present study are available upon reasonable request to the authors.

## ACKNOWLEDGMENTS

The authors would like to thank the participants, people with lived experience for their input during study planning and/or manuscript review (Lily Collison, Tommy Collison, Rachel Wobschall, Aleksys Patterson, Duncan Wyeth, June Kailes), the Cerebral Palsy Research Network for helping with recruitment and pre-review of the manuscript, and the Endowed Fund for Research in Cerebral Palsy Treatment for funding this study.

## Notes

### Competing Interest Statement

The authors have declared no competing interest.

### Funding Statement

This study was funded by the Endowed Fund for Research in Cerebral Palsy Treatment of Gillette Children's Specialty Healthcare.

### Author Declarations

The IRB of the University of Minnesota gave ethical approval for this work (STUDY00014717). All participants gave consent prior to participation.

